# Reference equations for peak oxygen uptake for treadmill cardiopulmonary exercise tests based on the NHANES lean body mass equations, a FRIEND registry study

**DOI:** 10.1101/2024.04.03.24305178

**Authors:** Everton J. Santana, Daniel Seung Kim, Jeffrey W Christle, Nicholas Cauwenberghs, Bettia E. Celestin, Jason V. Tso, Matthew T Wheeler, Euan A Ashley, James E. Peterman, Ross Arena, Matthew Harber, Leonard A. Kaminsky, Tatiana Kuznetsova, Jonathan N. Myers, Francois Haddad

## Abstract

**BACKGROUND:** Cardiorespiratory fitness (CRF), measured by peak oxygen uptake (VO_2_peak), is a strong predictor of mortality. Despite its widespread clinical use, current reference equations for VO_2_peak show distorted calibration in obese individuals. Using data from the Fitness Registry and the Importance of Exercise National Database (FRIEND), we sought to develop novel reference equations for VO_2_peak better calibrated for overweight/obese individuals - in both males and females, by considering body composition metrics.

**METHODS AND RESULTS:** Graded treadmill tests from 6,836 apparently healthy individuals were considered in data analysis. We used the National Health and Nutrition Examination Survey equations to estimate lean body mass (eLBM) and body fat percentage (eBF).

Multivariable regression was used to determine sex-specific equations for predicting VO_2_peak considering age terms, eLBM and eBF. The resultant equations were expressed as VO_2_peak (male) = 2633.4 + 48.7✕eLBM (kg) - 63.6✕eBF (%) - 0.23✕Age^2^ (R^2^=0.44) and VO_2_peak (female) = 1174.9 + 49.4✕eLBM (kg) - 21.7✕eBF (%) - 0.158✕Age^2^ (R^2^=0.53). These equations were well-calibrated in subgroups based on sex, age and body mass index (BMI), in contrast to the Wasserman equation. In addition, residuals for the percent-predicted VO_2_peak (ppVO_2_) were stable over the predicted VO_2_peak range, with low CRF defined as < 70% ppVO_2_ and average CRF defined between 85-115%.

**CONCLUSIONS:** The derived VO_2_peak reference equations provided physiologically explainable and were well-calibrated across the spectrum of age, sex and BMI. These equations will yield more accurate VO_2_peak evaluation, particularly in obese individuals.

## INTRODUCTION

Cardiorespiratory fitness (CRF) refers to the integrated capacity of the circulatory and respiratory systems to supply oxygen to skeletal muscle for energy production needed during physical activity (1–3). CRF, which is clinically evaluated during exercise testing as peak oxygen uptake (VO_2_peak), is established as one of the strongest biomarkers of health (4,5).

Population based studies have shown that several factors may influence VO_2_peak including age, sex, lean body mass (LBM), obesity, and physical activity level (6–11). To account for these differences, CRF is usually reported as relative value using percentage of predicted VO_2_peak (ppVO_2_, [measured VO_2_peak / predicted VO_2_peak] × 100%) (11–15). Historically, the most commonly used reference equation was developed by Hansen, Sue, and Wasserman (commonly termed the “Wasserman equation”) (16). The approach of Hansen *et al.* was innovative as it used a stepwise approach first to account for “ideal body weight” and then further adjusted for the presence of obesity or underweight status. While only derived in 77 male patients, the equation has been widely adopted. Due to methodological issues including the small sample size and overrepresentation of males in studies, the 2013 policy statement by the American Heart Association emphasized the need for improved reference standards of VO_2_peak (17).

Aiming at providing reference standards of VO_2_peak using directly measured CRF (rather than indirect estimates reported in previous studies) from a multicenter database with a representative sample of the United States population (17), the Fitness Registry and the Importance of Exercise: A National Database (FRIEND) registry was created. This has led to newly proposed reference equations for treadmill exercise testing, using either additive or multiplicative modeling based on mass, height or age terms ((12,14). While the Wasserman equations and prior FRIEND equations are extremely valuable, reference equations for VO_2_peak could be improved in some aspects. First, residual plot analyses revealed unequal bias in three groups: individuals with high VO_2_peak, females, and individuals with obesity (12).

Second, since the equations are often based on mass, they do not differentiate between the contributions of LBM or the decrease in CRF associated with obesity at a population level. Third, the equations may not always be well-calibrated for a general non-sedentary population. Finally, categorizations of CRF are most often cohort-specific and thus not always generalizable to a wider population.

For a more physiologic estimation of VO_2_peak we should consider LBM instead of total body mass. In fact, studies reported VO_2_peak as a function of LBM is a better predictor of clinical outcomes (18,19). Further adjustment for body fat percentage (BF) could be added to account for the potential decrease in CRF associated with obesity (20).

In this work, we aimed to develop better reference models that address the flaws of previous equations (**Figure 1**). To accomplish this, we leveraged the FRIEND (21,22)-(23) registry to develop two sets of reference equations based on estimated LBM (eLBM): 1) an equation for all the apparently healthy FRIEND participants adjusting for body fat, which helps to answer the question of whether CRF is reduced compared to individuals of matched body habitus; and 2) a healthy weight group in order to assess how fit a given individual generally is. We subsequently tested the equations to assess if they were well calibrated across sex, age, and BMI subgroups. We then determined whether the novel equation using eLBM and body fat improved the detection of HF as compared to the Wasserman equation.

**Figure 1.**
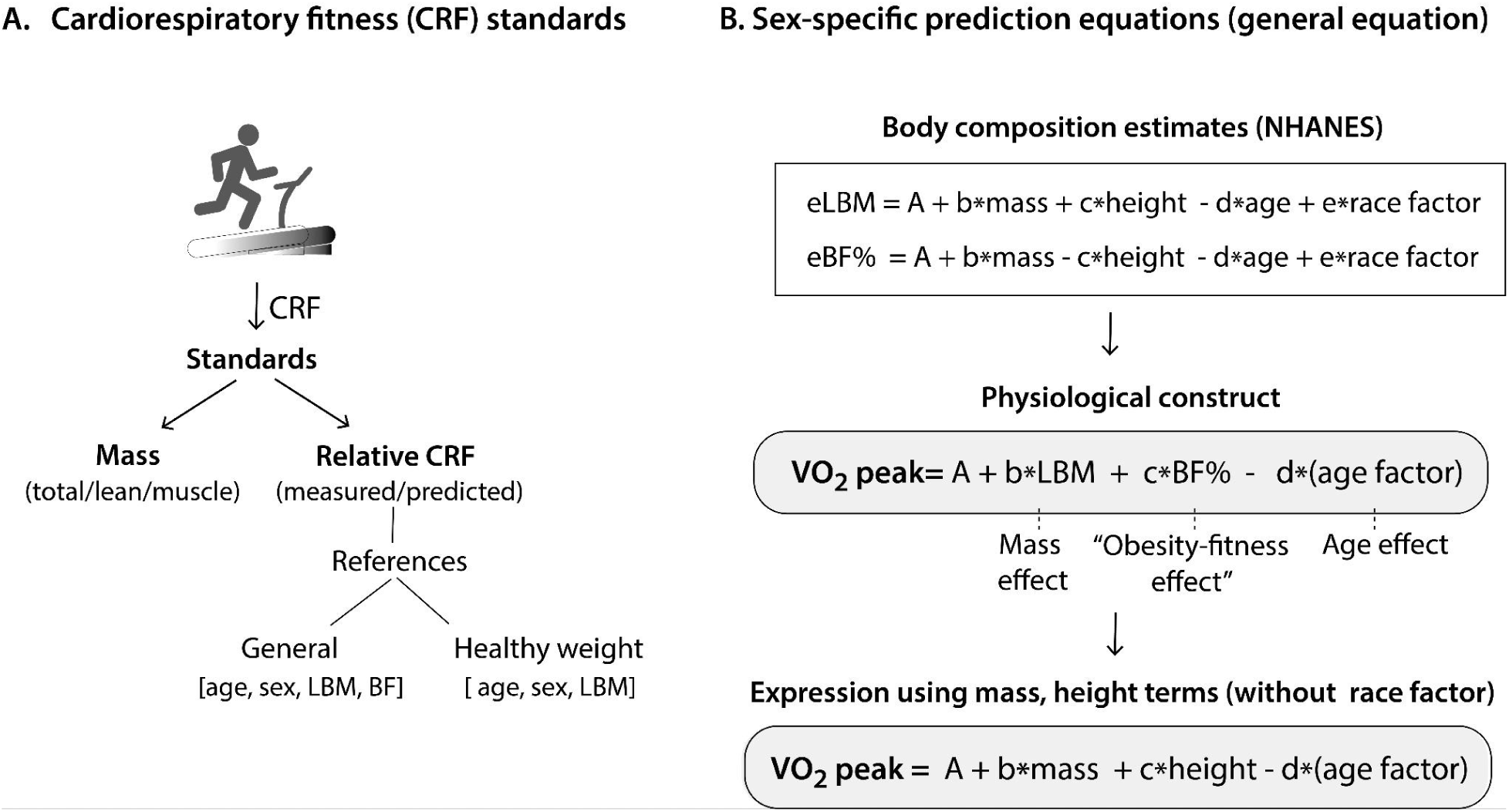
Study rationale and derived equations for percent-predicted VO_2_peak from the FRIEND data. In this study, the “obesity-fitness effect” refers to the usual decrease in cardio-respiratory fitness associated with obesity in cohort-based studies.

## METHODS

### Study cohort

The FRIEND registry is a multi-institutional initiative established in 2014 with the primary goal of developing normative CRF values for the United States across the adult lifespan. Geographical representation in the FRIEND registry included one or more tests from all 50 states apart from Alaska, Nebraska, and Wyoming. In this study, we analyzed graded treadmill Cardiopulmonary exercise test (CPX) tests. All centers participating in FRIEND had individual institutional review board approval. The procedures used in acquiring and managing the FRIEND registry data have been previously reported (17,24). Briefly, all CPX centers contributing data were experienced in CPX testing and used valid and reliable calibration and testing procedures consistent with prior guideline-based recommendations (25). Data collection for FRIEND included demographics, anthropomorphic information (height, mass, and BMI), vital signs, and major medical comorbidities. During the CPX tests, VO_2_peak was directly measured by open-circuit spirometry (mL O_2_ ⋅ kg^-1^ ⋅ min^-1^) and peak heart rate, treadmill speed, and treadmill fractional grade were also recorded.

#### Apparently healthy participant selection

We selected apparently healthy participants that underwent a graded treadmill exercise at participating centers of the FRIEND registry. We excluded participants who underwent a non-graded treadmill CPX, had a peak respiratory exchange ratio (RER) <1.0, age ≥ 80 years, BMI (kg⋅m^-2^) < 18.5 or ≥ 40 (groups with low sample size), and other quality criteria including low peak heart rate or non-physiological outlier values (see **Supplemental Methods**).

The definition of apparently healthy participants (**Figure 2**) excluded those who had cardiovascular disease, chronic kidney disease, chronic lung disease, endocrine or neurological disorders or diabetes mellitus. Participants with test indications for symptoms (dyspnea, chest pain, shortness of breath, exercise intolerance or systolic function) or electrocardiographic abnormalities were also excluded. Participants with hypertension or dyslipidemia were not excluded, consistent with prior fitness studies or registries (26). Hypertension was defined as either a resting blood pressure >140/90 mmHg or a medically documented diagnosis. We further excluded outliers using median absolute deviation (with a deviation criterion of 3) in subgroups split by sex and BMI considering the ratio between VO_2_peak and workload. A total of 6,836 apparently healthy participants with BMI between 18-25 and not on beta-blockers, remained for analysis.

**Figure 2.**
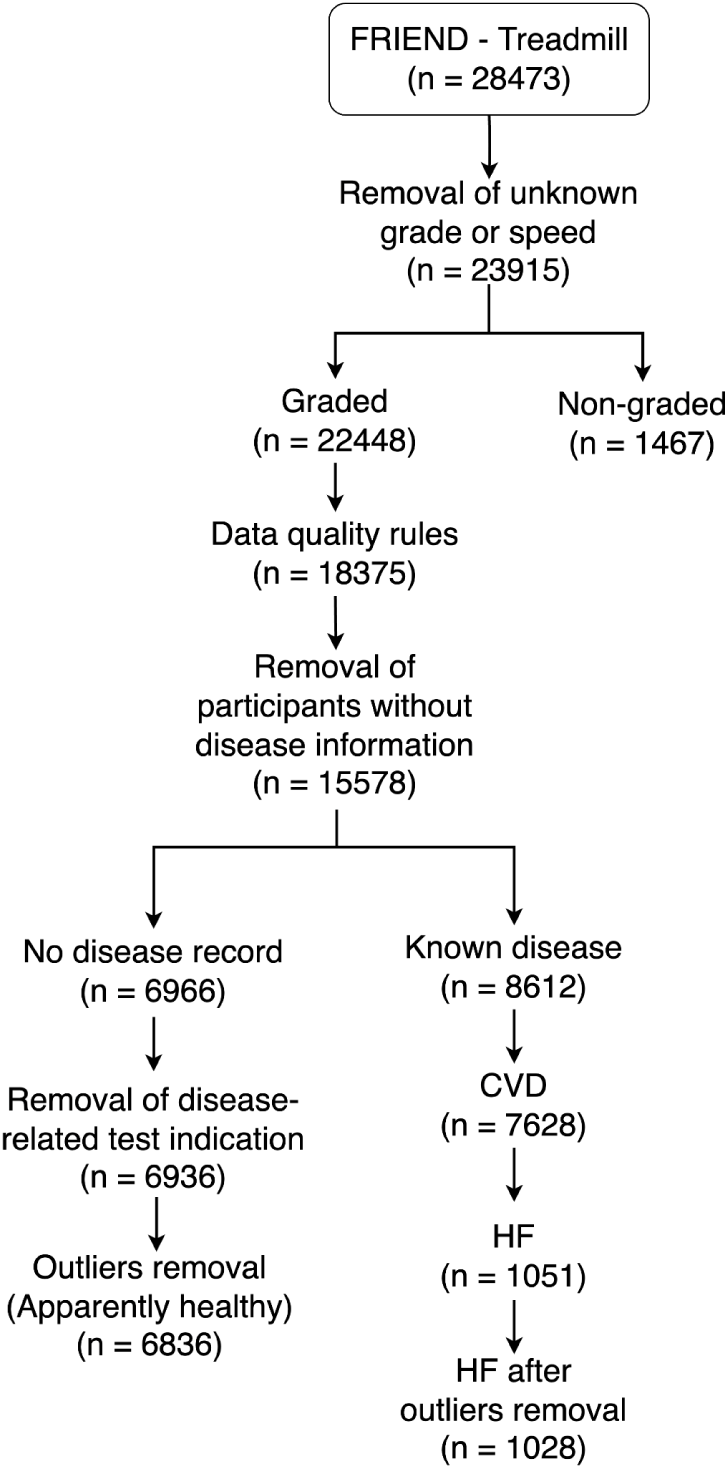
FRIEND consort diagram.

### Patients with heart failure

A subgroup of patients from the FRIEND registry who had a diagnosis of cardiomyopathy or heart failure (HF) were included in the HF subgroup (n=1028). This group was added to the analysis to evaluate the ppVO_2_ calculated with the novel reference equations to detect HF.

### Rationale for using additive sex-specific models for predicting VO_2_peak

In this study, we adopted an additive model for absolute VO_2_peak with eLBM, estimated BF (eBF) and age^2^ terms as dependent variables. By choosing an additive model, the eLBM and eBF terms can then be simplified to mass, height and age terms using the coefficients of the NHANES formulas (27) without requiring first to calculate eLBM and eBF. We selected sex-specific equations for the additive models to avoid introducing several interaction terms in the equation.

### Statistical analysis

Python 3.11 was used for statistical analyses. Multivariable linear regression was used to generate equations for predicted VO_2_peak. Data were summarized as mean and standard deviation (SD) for continuous variables and percentages for categorical variables.

#### Radar plots to visualize calibration of previous equations in subgroups

Radar plots of ppVO_2_ values according to the FRIEND and Wasserman equations were presented by sex according to medians in three age (<40, 40-60, > 60 years) and three BMI subgroups (18.5-24.9, 25-29.9 and ≥30 kg^-^m^-2^).

#### Novel equation development

Reference equations were derived in the apparently healthy cohort (N=6,836). In this step, we considered the variables eLBM, eBF and age^2^. Since direct measures of LBM and body fat via Dual-electron X-ray Absorptiometry (DXA) are not typically available in clinical practice, we used the validated NHANES equations (**Figure 1**) (27). eBF was used to estimate the “obesity-fitness effect” or the decrease in CRF often associated with obesity at a population level (20,28–30). We assessed whether ppVO_2_ yielded equal variance across the VO_2_peak range and proposed thresholds for CRF categories based on the ppVO_2_ distribution.

To allow comparisons to a reference healthy weight group, we also developed equations for predicting VO_2_peak in the apparently healthy participants that were in the normal BMI weight category, i.e., 18.5 ≤BMI <25 kg⋅m^-2^ (N=1,693 males and N=1,514 females). For this, eLBM and age^2^ were considered as input variables.

#### Comparison of equations for identification of prevalent heart failure

Using the HF subset (N=1,028) in conjunction with the apparently healthy cohort (N=6,836), we compared our novel equation and the Wasserman equation in their ability to classify HF by comparing the thresholds of both equations for the 5^th^ percentile (low CRF threshold), stratified by sex. Using data from the contingency tables, we compared the cumulative frequency plots of the two equations for identifying HF patients stratified by sex and calculated the diagnostic odds ratio (DOR) (31). To compare the DORs, we calculated the standard error based on the elements of the 2×2 contingency table and analyzed the difference in significance using two-tailed z-tests (32).

## RESULTS

### Characteristics of the analyzed FRIEND subset

A flow diagram summarizing the selection of the analyzed subset of FRIEND is presented in **Figure 2**. In brief, the FRIEND dataset analyzed was from the April 2021 data freeze. Of the 15,578 participants with linked disease phenotype data who underwent CPX testing with a graded treadmill protocol, 8,612 participants were excluded due to known disease. After removal of outliers and disease-related test indications, a total of 6,836 participants with CPX tests remained for analysis as an apparently healthy sample. Among the patients with disease, 7,628 (88.6%) had cardiovascular disease, of which a subset of 1,028 had known HF.

Of the apparently healthy subset (N=6,836), the average age was 44±14 years and 43.4% (N=2,968) were female. Men were on average taller (1.79±0.07 vs 1.65±0.06 meters), weighed more (84±14 vs 71±14 kg), had greater eLBM (58.9±7.4 vs 41.2±5.6 kg), and had lower eBF (26.7±4.2 vs 37.5±5.1 %). The HF group (N=1,028) was generally older (52±15 vs 44±14 years) and achieved lower maximal heart rate (139±27 vs 175±16 beats per minute), % predicted heart rate (82.7±13.5 vs 100.0±7.4%), and VO_2_peak (23.7±.5 vs 35.9±11.1 mL⋅min^-1^⋅kg^-1^). These data are summarized in **Table 1**.

**Table 1.** Clinical characteristics of the FRIEND subcohorts presented as mean ± standard deviation. DBP: diastolic blood pressure; HR: heart rate; RER: respiratory exchange ratio; SBP: systolic blood pressure.

|  | Apparently healthy |  |  | Heart failure |  |  |
| --- | --- | --- | --- | --- | --- | --- |
| Characteristic | Total<br>(n=6836) | Female<br>(n=2968) | Male<br>(n=3868) | Total<br>(n=1028) | Female<br>(n=334) | Male<br>(n=694) |
| **Anthropometrics** |  |  |  |  |  |  |
| Age (years) | 44 ± 14 | 45 ± 14 | 43 ± 14 | 52 ± 15 | 50 ± 14 | 53 ± 15 |
| Height (m) | 1.72 ± 0.10 | 1.65 ± 0.06 | 1.79 ± 0.07 | 1.73 ± 0.10 | 1.64 ± 0.07 | 1.77 ± 0.08 |
| Weight (kg) | 78 ± 16 | 71 ± 14 | 84 ± 14 | 83 ± 17 | 73 ± 15 | 88 ± 16 |
| BMI (kg·m<sup>-2</sup>) | 26 ± 4 | 26 ± 5 | 26 ± 4 | 28 ± 5 | 27 ± 5 | 28 ± 4 |
| eLBM (kg) | 51.2 ± 11.0 | 41.2 ± 5.6 | 58.9 ± 7.4 | 54.2 ± 11.5 | 42.0 ± 5.9 | 60.0 ± 8.5 |
| eBF (%) | 31.4 ± 7.1 | 37.5 ± 5.1 | 26.7 ± 4.2 | 32.2 ± 6.6 | 38.9 ± 5.3 | 28.9 ± 4.4 |
| **Baseline vitals and test measurements** |  |  |  |  |  |  |
| Resting SBP (mmHg) | 120 ± 14 | 115 ± 14 | 124 ± 13 | 116 ± 20 | 119 ± 21 | 115 ± 19 |
| Resting DBP (mmHg) | 77 ± 10 | 74 ± 10 | 80 ± 9 | 72 ± 12 | 73 ± 11 | 72 ± 12 |
| Max HR (bpm) | 175 ± 16 | 174 ± 17 | 177 ± 16 | 139 ± 27 | 142 ± 26 | 138 ± 28 |
| % Predicted HR (%) | 100.0 ± 7.4 | 99.5 ± 7.7 | 100.3 ± 7.2 | 82.7 ± 13.5 | 83.4 ± 13.2 | 82.3 ± 13.6 |
| Max RER | 1.18 ± 0.10 | 1.17 ± 0.10 | 1.18 ± 0.10 | 1.14 ± 0.10 | 1.12 ± 0.09 | 1.15 ± 0.10 |
| VO<sub>2</sub>peak (mL·min<sup>-1</sup>) | 2775.8 ± 924.9 | 2048.8 ± 521.6 | 3333.6 ± 765.7 | 1947.9 ± 820.1 | 1526.6 ± 500.1 | 2150.7 ± 866.0 |
| VO<sub>2</sub>peak/Mass (mL·kg<sup>-1</sup>·min<sup>-1</sup>) | 35.9 ± 11.1 | 29.8 ± 8.5 | 40.6 ± 10.6 | 23.7 ± 9.5 | 21.2 ± 7.0 | 24.8 ± 10.3 |

### The calibration of previous FRIEND and Wasserman equations across age, sex and BMI

Prior to deriving new reference equations, we visualized the calibration of prior equations (12,14,16) using radar plots by means of median ppVO_2_ in subgroups stratified according to sex, age and BMI. Overall, the previously published FRIEND equations were better calibrated across all age and BMI groups in males than in females. In females, these equations notably overestimated ppVO_2_ in the obese groups between 40-60 and >60 years of age (**Figure 3A).** The Wasserman equation slightly over-estimated ppVO_2_ in nearly any subgroups in males but overestimated ppVO_2_ in all BMI and age groups in females (**Figure 3B**).

**Figure 3.**
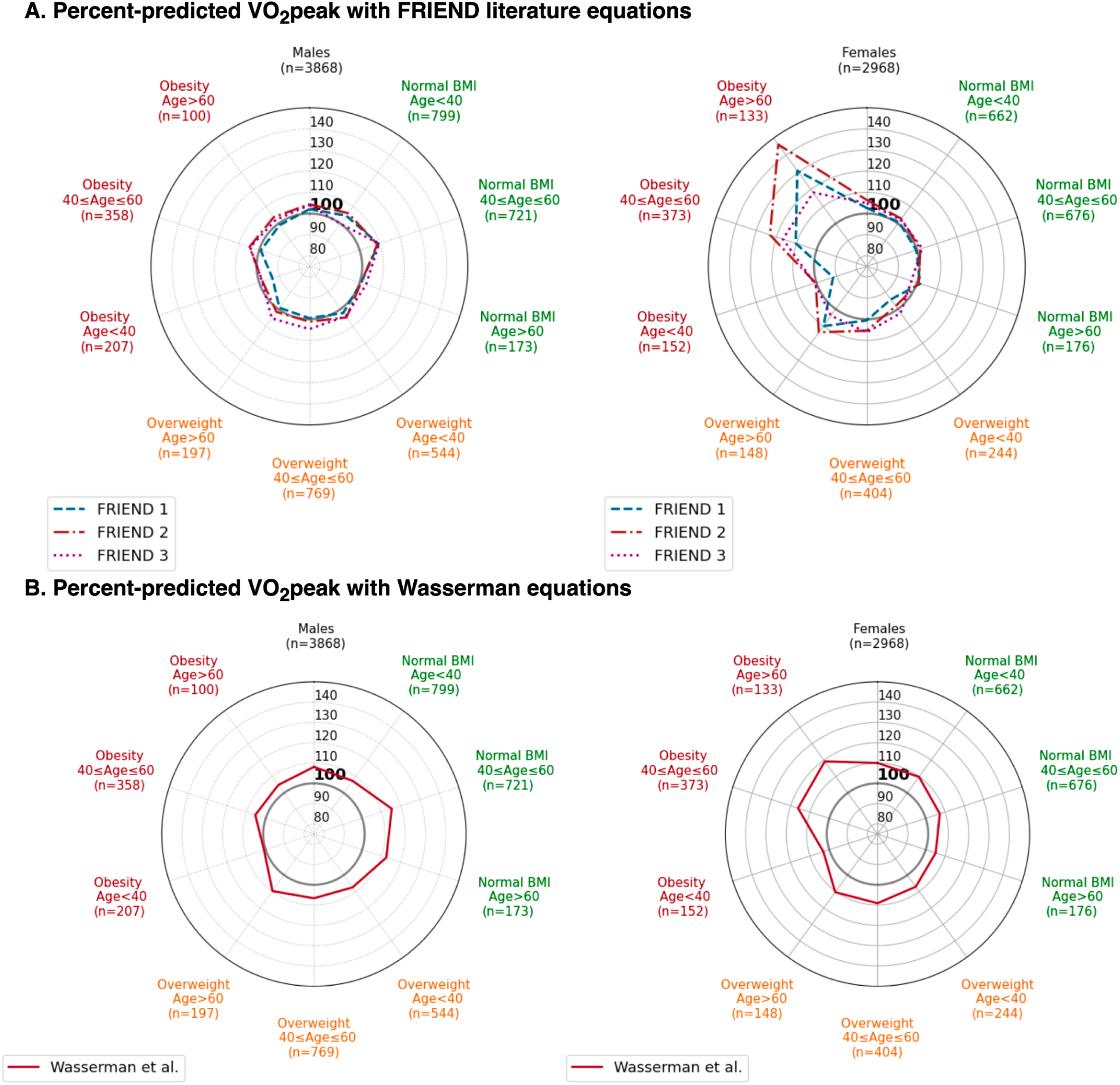
Comparison of equations demonstrates overestimation, particularly in obese females, in both FRIEND and Wasserman equations. FRIEND 1: Myers *et al*(*12*); FRIEND 2: Silva *et al*(*11*), FRIEND 3: Nevill *et al*(*14*).

#### Novel reference equations for VO_2_peak and percent-predicted VO_2_peak

We developed two sets of sex-specific equations: one considering the entire apparently healthy FRIEND data across the spectrum of BMI (N=6,836) and another for a healthy weight reference population (N=3,207).

For the apparently healthy cohort, the reference equation for males was VO_2_peak = 2633.4 + 48.7*eLBM (kg) - 63.6*eBF (%) - 0.23*age^2^ (R^2^=0.44) and for females, VO_2_peak = 1174.9 + 49.4*eLBM (kg) - 21.7*eBF (%) - 0.16*age^2^ (R^2^=0.53). For the healthy weight cohort, the reference equation for males (N=1,693) was VO_2_peak = 1298.8 + 48.3*eLBM (kg) - 0.30*age^2^ (R^2^=0.41), while for females (N=1,514) the derived equation was VO_2_peak = 485.2 + 50.7*eLBM (kg) - 0.20* age^2^ (R^2^=0.51). These are summarized in **Table 2** along with the simple transformation to mass and height terms using the NHANES equations. As shown in **Supplemental Figure 1**, the measured versus predicted VO_2_peak had a slope equal to 1 for all equations.

**Table 2.**
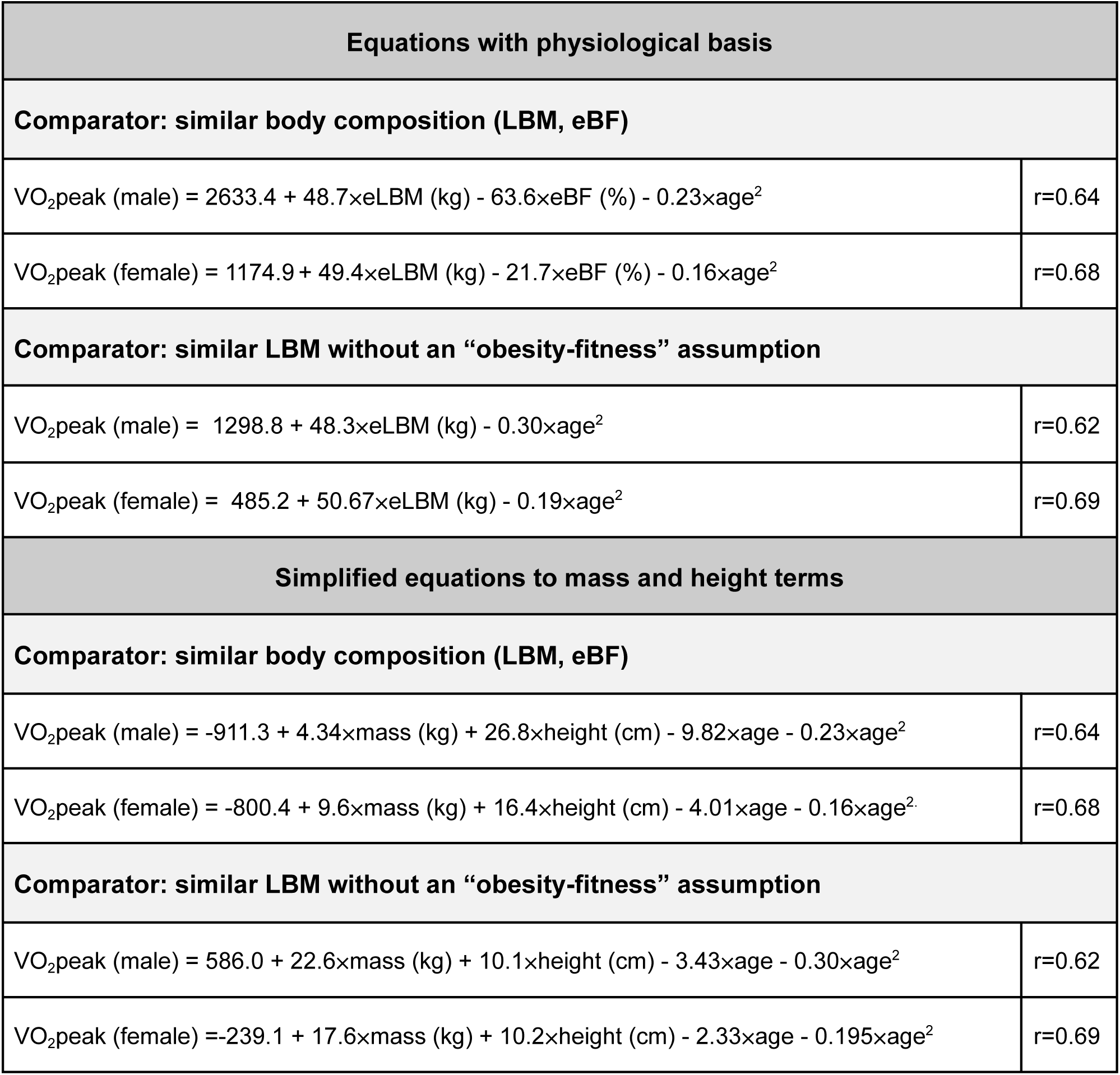
Reference equations for VO_2_peak (mL·min^-1^) developed in this study.

In the apparently healthy cohort, the median ppVO2 was 100.2% in males and 99.0% in females. **Figure 4A-B** shows the percent-predicted versus predicted VO_2_peak for males and females, respectively. We tested, using quantile regression, whether ppVO_2_ was stable (constant) across the VO_2_peak predicted range. For male and females, the pseudo R^2^ explained less than 1% of the variance for quantile functions at 5, 20, 80, 95^th^ percentiles suggesting quantile specific stability of variance.

**Figure 4.**
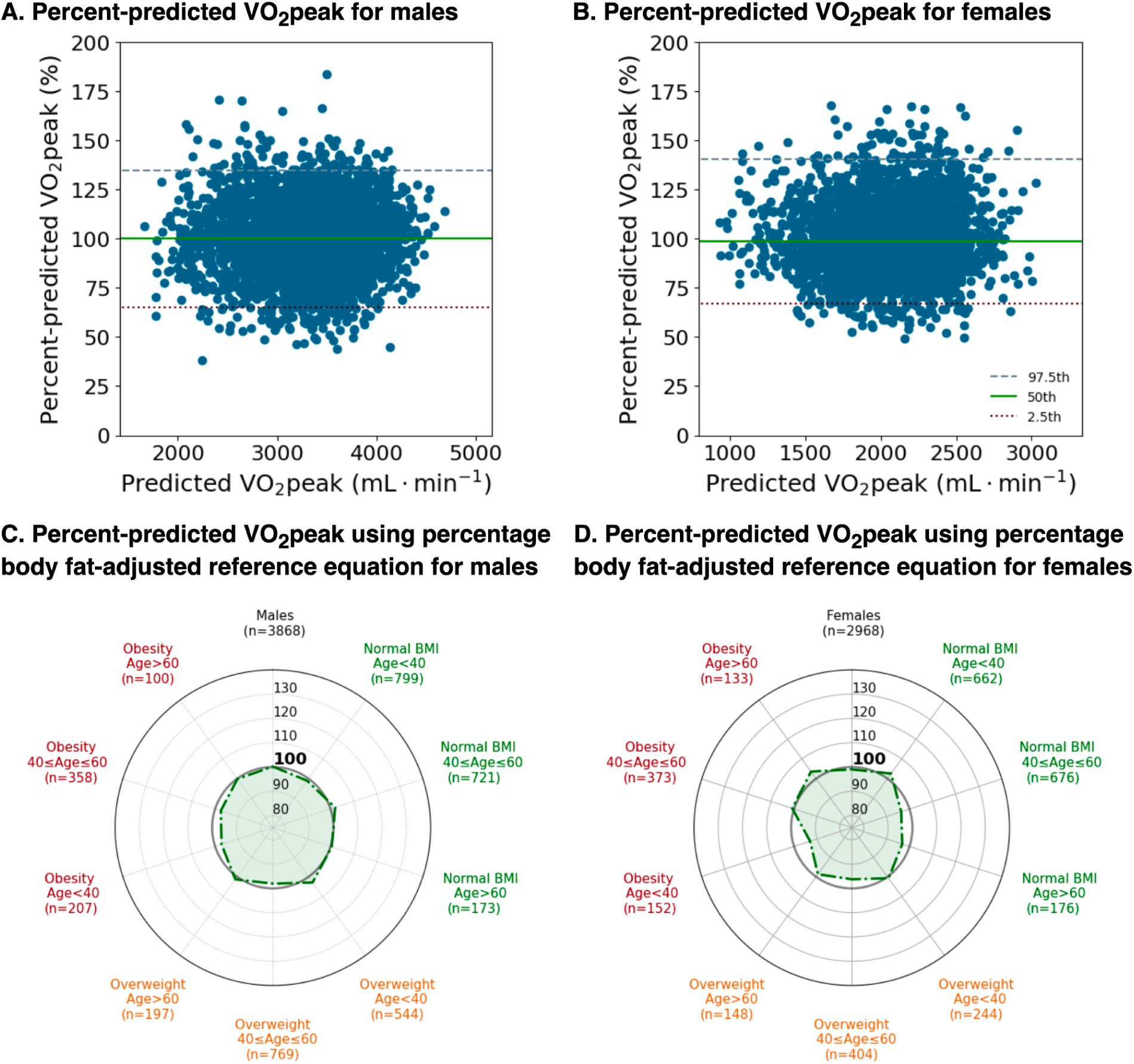
Assessment of using percentage body fat-adjusted reference equation for percent-predicted VO_2_peak.

We then examined the median ppVO_2_ across the subgroups according to sex, age and BMI. Radar plots (**Figure 4C-D)** demonstrated that all subgroups were overall well-calibrated, with median ppVO_2_ within the range 97-103% for males and 93-104% for females. The greatest improvement compared to the Wasserman equation was observed for the females older than 60 with obesity (median ppVO_2_ from 119.2 to 103.5, p<0.001 for Mann-Whitney U test).

To determine categories of CRF, we first analyzed the percentile distribution of ppVO_2_ for males and females (**Figure 5**). In choosing the representative thresholds, we opted for a simple and concordant classification for males and females. The 2.5^th^ to 5^th^ percentile of ppVO_2_ was approximately 70% (71.5% in males and 73% in females), indicating this as the threshold for low CRF; the ppVO_2_ of 85-115% corresponded to the 20^th^ to 80^th^ percentile and was stable for both male and female; for the high CRF threshold, we chose a ppVO_2_ of 135%, corresponding to a percentile of 97.5. The reference equations derived in the healthy weight cohort presented similar ppVO_2_ for the considered thresholds.

**Figure 5.**
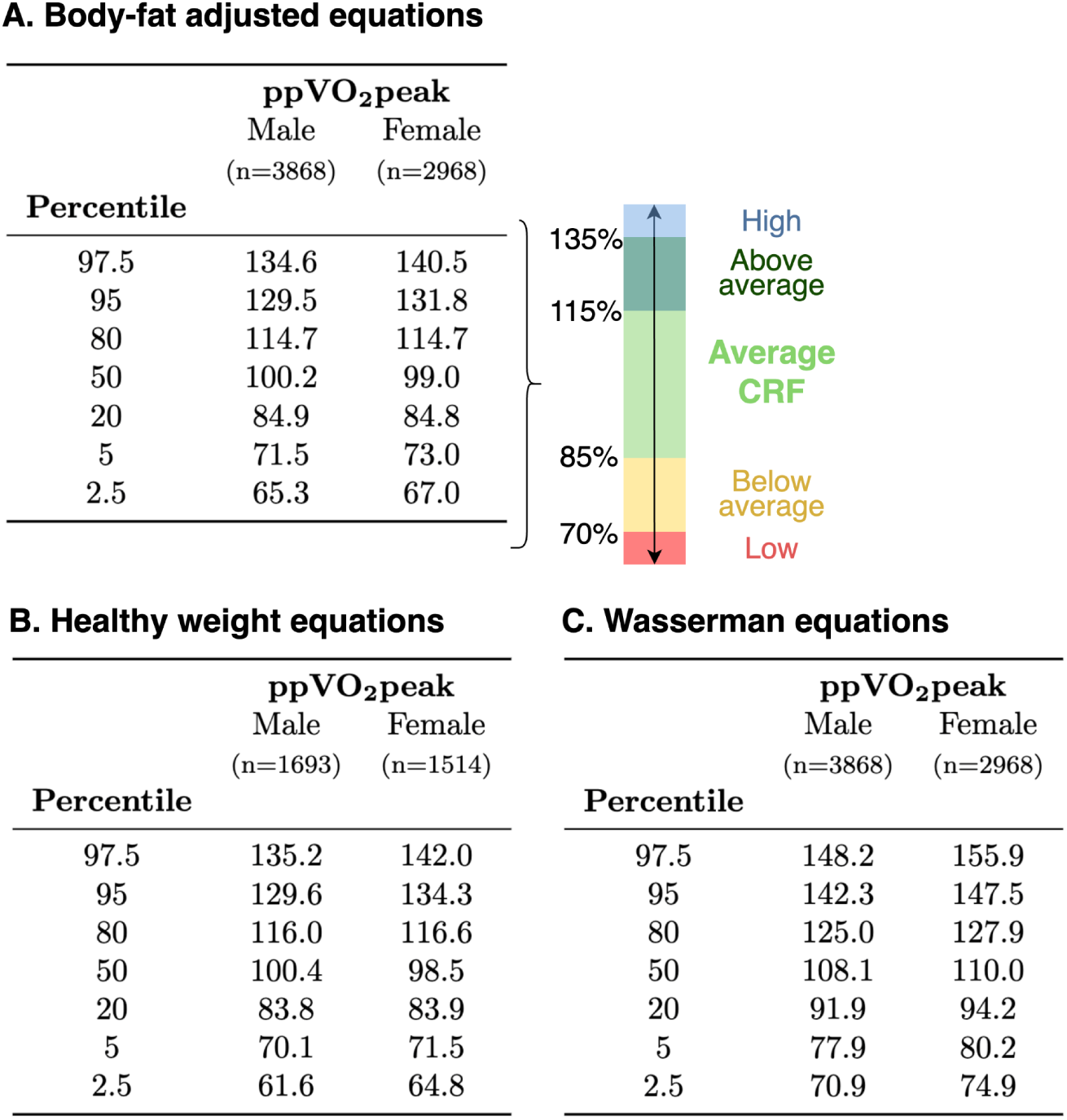
Percentiles and percent-predicted VO_2_peak-based cardiorespiratory fitness categories thresholds.

We also presented percentile distribution in parallel for the Wasserman equation. Considering the 5^th^ percentile the low CRF category threshold, the equivalent ppVO_2_ using Wasserman corresponded to 80% (77.9% in males and 80.2% in females). Wasserman and colleagues previously recommended a ppVO_2_ threshold of 83% for identifying prevalent HF (16).

#### Quantifying the differences in the effects of obesity in the apparently healthy cohort

The effects of obesity in our cohort was not stable across the spectrum of BMI in both males and females using the different set of equations. We quantified the decrease in CRF in the groups stratified by sex and BMI by comparing the ppVO_2_ using the reference equation in the healthy weight group with ppVO_2_ using the eBF-adjusted equation (**Table 3**). In males, the decrease was on average 7% in the overweight group and 15% in individuals with obesity. For females, the decrease was in 5 and 10% for overweight and obesity, respectively.

**Table 3.** Relative difference(%) in percent-predicted VO_2_peak (%) calculated using the equation derived in the healthy weight group (HW, BMI 18-25) in relation to the body fat-adjusted equation.

|  | **Healthy weight**<br>(n=1693) | **Overweight**<br>(n=1510) | **Obesity**<br>(n=665) |
| --- | --- | --- | --- |
| Male | 0 ± 2 | -7 ± 2* | -15 ± 3^ |
| Female | 0 ± 2 | -5 ± 1* | -10 ± 1^ |

Relative difference was assessed using the formula $[\text{ppVO}_2(\text{HW}) - \text{ppVO}_2(\text{eBF})] / \text{ppVO}_2(\text{HW})$ .
\* p<0.001 for two-sided t-test between Healthy weight and Overweight;
^ p<0.001 for two-sided t-test between Healthy weight and Obesity.

#### Discrimination of HF by previous and novel equations for ppVO2

Using the subset of FRIEND with known HF (N=1,028), the cumulative frequency plots are presented for ppVO_2_ using the Wasserman and the novel FRIEND equations and the diagnostic odds for the diagnosis of HF. As expected, the frequency plots using the Wasserman equations were shifted to higher values (**Figure 6A and C**). In the apparently healthy weight cohort, the median ppVO_2_ was shifted to the right with 8% higher median in males and 11% in females. In the HF cohort, the median ppVO_2_ using Wasserman was 5% higher in males and 9% in females. In the obesity group, a similar shift pattern was observed (**Supplemental Figure 2**). Based on these data, we present suggested ppVO_2_ thresholds for categories of CRF in **Supplemental Table 1.**

**Figure 6.**
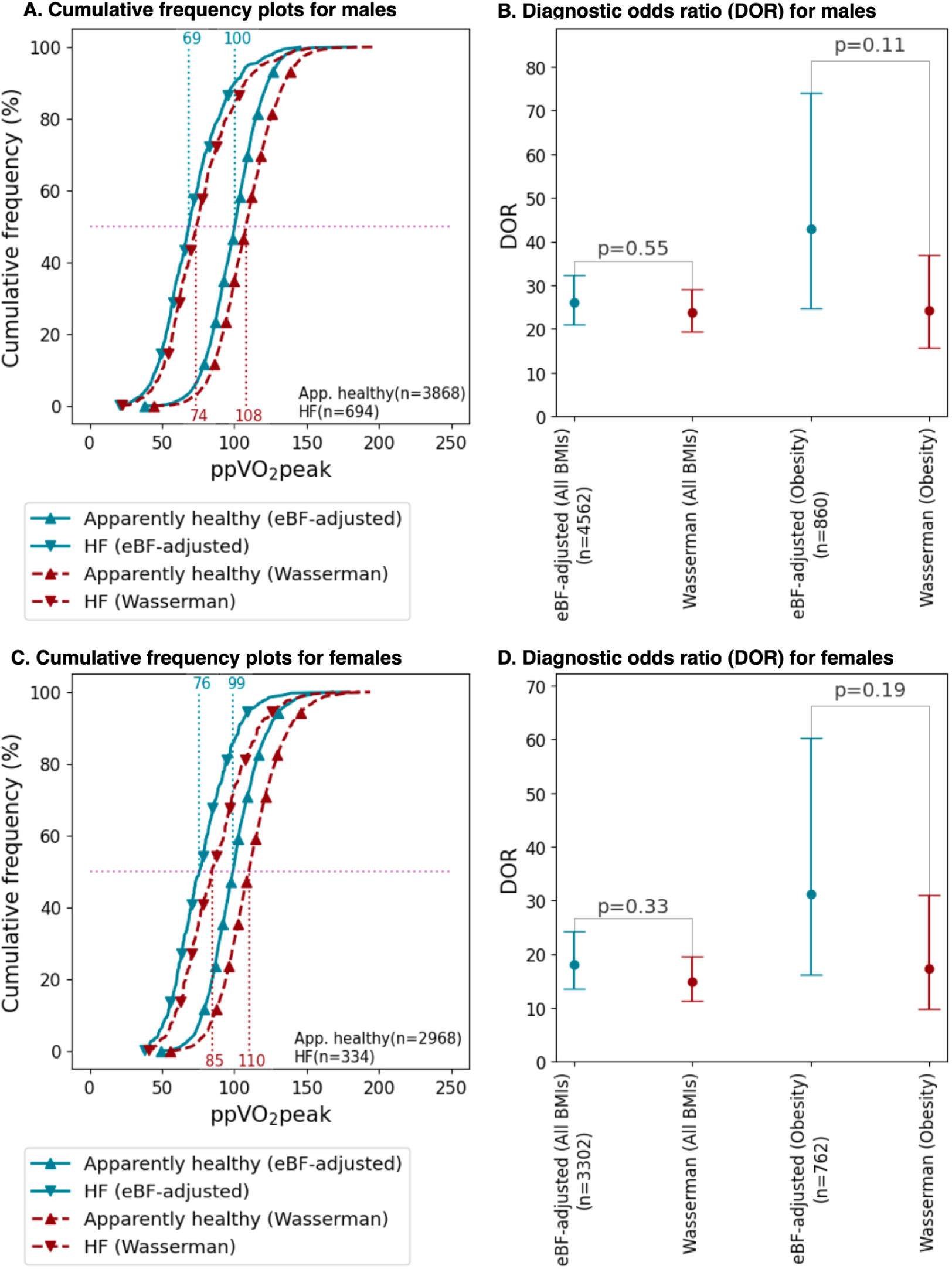
Application of novel eBF-adjusted VO_2_peak equation in diagnosing heart failure (HF), as compared to the Wasserman equation.

The diagnostic odds ratios for the diagnosis of HF (FRIEND vs. Wasserman) for males was 26 vs. 24 (*P*=0.55) in the entire group vs. 42.8 vs. 24.2 (*P*=0.11) in the subset with obesity (N=860, **Figure 6B**). In females, the diagnosis of HF was 18 vs. 15 (*P*=0.33) in the entire group vs. 31.2 vs. 17.4 (*P*=0.19) in the subset with obesity (N=762, **Figure 6D**).

## DISCUSSION

Our study has three main findings. First, novel sex-specific reference equations for CRF that build on the physiological construct of LBM demonstrated improved calibration among subgroups stratified according to sex, age and BMI (**Figures 3** and **4**). Second, we developed two sets of reference equations that allow reporting of CRF with or without correction for the “obesity-fitness effect”. In other CRF equations, this information is often embedded in the equations. Finally, a grading system for CRF derived from ppVO2 percentile distributions may help standardize reporting of CRF in clinical practice when using either the FRIEND or the Wasserman equation.

Since CRF varies according to age, sex, body composition, and physical activity levels, reporting of CRF requires the development of well-calibrated prediction equations. Despite being developed in a cohort of only 77 male shipyard workers, the Wasserman equation remains the most used in clinical practice. It has endured the test of time, since it was based on sound physiological considerations. In fact, Wasserman developed a stepwise approach for estimating VO_2_peak first considering the contribution of ideal body weight and then correcting for the “excess mass” or underweight status. This was chosen since fat mass does not contribute equally to VO_2_ during exercise. Ideal body mass was a concept developed for actuarial survival by insurance companies. Since then, LBM has emerged as a measure of “energetic mass” during physical activity (33). For example, Krachler *et al*. reported from the DR’s Extra study that scaling to LBM led to body size independent scaling of VO_2_peak compared to scaling to total mass which led to underestimation of CRF in obesity (18). The meta-analysis by Lolli *et al*. on 6,514 participants also confirmed that the allometric coefficient for scaling VO_2_peak based on LBM is close to unity (34).

In our study, we developed novel equations for CRF that build on the physiological construct of LBM but that can be easily simplified to mass and height terms. We derived sex-specific equations not only to account for different rates of decline of CRF but also for potential differences of excess weight on CRF. In our equations, the eLBM constant was close to 50 in both males and females, consistent with the constant identified by Batterhan *et al.*, Krachler *et al.*, and Lorenzo *et al*. (18,33,35) Moreover, the LBM constant was stable in both the general and normal weight equations. Our study also confirmed that at a cohort based level, excess “weight” is associated with an average decreased CRF. This was also observed in the study of Jackson *et. al*. based on the Aerobics Center Longitudinal Study (ACLS) (36), the veterans based exercise study of Souza de Silva *et al.* (*37*). and the small observational study in adolescents of Ekelund *et al.* (*38*). Consistent with the study of Jackson et. al. we also observed a greater decrease in VO_2_peak with excess body fat in males and females (36). However, the effects of obesity on CRF may vary according to the physical activity level of an individual. For example, in the ACLS study, more sedentary individuals had an average lower achieved MET level on treadmill exercise testing (36). Clinicians may want to report relative CRF independent of an “obesity-fitness” effect. This was the impetus in our study to develop normal weight equations. We also quantified the embedded assumption of the obesity-fitness effect with an average decrease of CRF in obesity of 15 and 10% in males and females respectively. Consistent with the prior studies of Fleg *et al*. and Weiss *et al*., we have also observed a different sex-rate of decline in CRF (39). We have also used polynomial modeling of age to better reflect the non-linear decline of CRF with age in cross-sectional cohort based studies.

Our study also highlights the importance of considering subgroup calibration and stable variance or relative reference metric across the predicted range. While the prior FRIEND equations were all well-calibrated in the overall cohort, sub-group analysis demonstrated that calibration was needed in females with obesity (11,12,14). This subgroup bias is particularly important as older females with obesity are often referred for investigation of unexplained dyspnea (40). This subgroup bias is most likely the consequence of deriving a common equation for males and females, which neglects sex-age and sex-obesity interactions. In addition, calibration in obesity may be particularly poor when deriving equations based on total body weight standards, which are known to underestimate CRF in obesity (34). Using linear age terms can also underestimate CRF in older individuals and can further underestimate CRF in obesity. As demonstrated by Nevill *et al.*, allometric or multiplicative modeling improves calibration which could have been further improved by using sex-specific equations (14). Nevill et al. have also shown that the allometric coefficients for mass and height embed information on both LBM and the influence of obesity on CRF (14). In our study, we revealed these implicit assumptions and provide equations that allow reporting of relative CRF with or without accounting for population based obesity-fitness effect.

In clinical practice, there is variable reporting of categories of CRF. To use ppVO_2_ as a metric for reporting, stability in variance needs to be demonstrated across the predicted range. Using quantile regression analysis of variance, we have shown that ppVO_2_ is a simple and appropriate metric for reporting CRF. Based on percentile distribution, we proposed five ppVO_2_-based category thresholds of CRF which were consistent for both the normal weight and general reference cohorts (see **Supplemental Table 1**). The median value for the Wasserman et al. equation of 110% also highlights the fact that it is representative of an overall sedentary cohort. We also confirmed that 80% ppVO_2_ using the Wasserman equation corresponded to low CRF in the FRIEND cohort, though we note that this finding is based on percentile distribution, not outcomes. Outcome based studies have further developed grading systems especially in individuals with low CRF; 50% ppVO_2_ has often been reported as a threshold for severe dysfunction in HF for example (13,41). Further work will be needed to determine the threshold of ppVO_2_ that best predicts adverse cardiovascular events and classification with various disease states (e.g., heart failure).

In our HF sub-cohort analysis, we demonstrated that ppVO_2_ using the updated FRIEND or Wasserman equation had a strong diagnostic odds ratio for the diagnosis of HF. While the updated FRIEND equation has a nominally higher DOR than the Wasserman equation, the cohort was underpowered to show differences in obesity. This will require further investigation focused on classification of disease and outcome analysis.

Several limitations in our study should be noted. First, we estimated LBM and BF using the NHANES equation (27). The NHANES equations were however derived from large cohorts and were internally validated. Second, our study was mainly representative of a White race and future studies will be needed in diverse populations. Third, while all FRIEND registry sites followed current guidelines, there was variability in the choice of treadmill protocols, equipment, and data collection procedures. Fourth, while the reference equations are based upon broad recruitment of both sexes from the FRIEND registry, cohort characteristics, such as underlying physical activity levels, may affect its generalizability and the obesity-fitness effect noted in our work. Finally, future outcome studies with a greater focus on females, individuals with obesity or the elderly will be needed to understand the added diagnostic and prognostic value of the novel equations.

In conclusion, we developed well calibrated and explainable equations for peak VO_2_ during treadmill exercise testing. This will likely help standardize and reduce bias in clinical reporting.

## Acknowledgements

We thank all FRIEND study subjects for their participation in this research. FRIEND Consortium Contributors are as follows: Ball State University (Leonard Kaminsky, Matthew Harber, Mitchell Whaley), Biodynamics and Human Performance Center, Georgia Southern University Armstrong Campus (Gregory Grosicki, Andrew Flatt, and Meral Culver), Brooke Army Medical Center (Kenneth Leclerc), Clay Exercise Science Center at the University of Mount Union (Katherine Clark), Cone Health (Paul Chase), Cleveland Clinic (John Kirwan), Fitness Institute of Texas in the Department of Kinesiology and Health Education at the University of Texas at Austin (Philip Stanford), Goshen Heart and Vascular Center (Matthew Thomas), Johns Hopkins University (Kerry Stewart), Longwood University (Jo Morrison), North Carolina Wesleyan College (Meir Magal), Pepperdine University, Saint Francis University (Kristofer Wisniewski), San Francisco State University (Jimmy Bagley), Southern Connecticut State University (Robert Axtell), Taylor University (Erik Hayes), University of Kansas Medical Center (Sandy Billinger), University of Montana (Charles Dumke and Katherine Christison), VA Palo Alto Health Care System (Jonathan Myers).

## Data Availability

Data will be available to qualified investigators upon request to the FRIEND steering committee.

## Supplemental Methods

Exclusion conditions:

- Peak respiratory exchange ratio < 1 or ≥ 1.75
- Body mass index (kg·m^-2^) < 18.5 or ≥ 40
- Age (years) ≥ 80
- Height (m) < 1.5 or > 2.2
- Weight (kg) < 40 or > 150
- Fractional grade = 0 or > 28
- Maximal heart rate (bpm) < 60 or > 250
- Body surface area (m^2^) > 2.7

Disproportional relations between speed (m·min^-1^) and VO_2_peak (in METs) were also excluded (54 individuals in total).

## Supplemental Figures

**Supplemental Figure 1.**
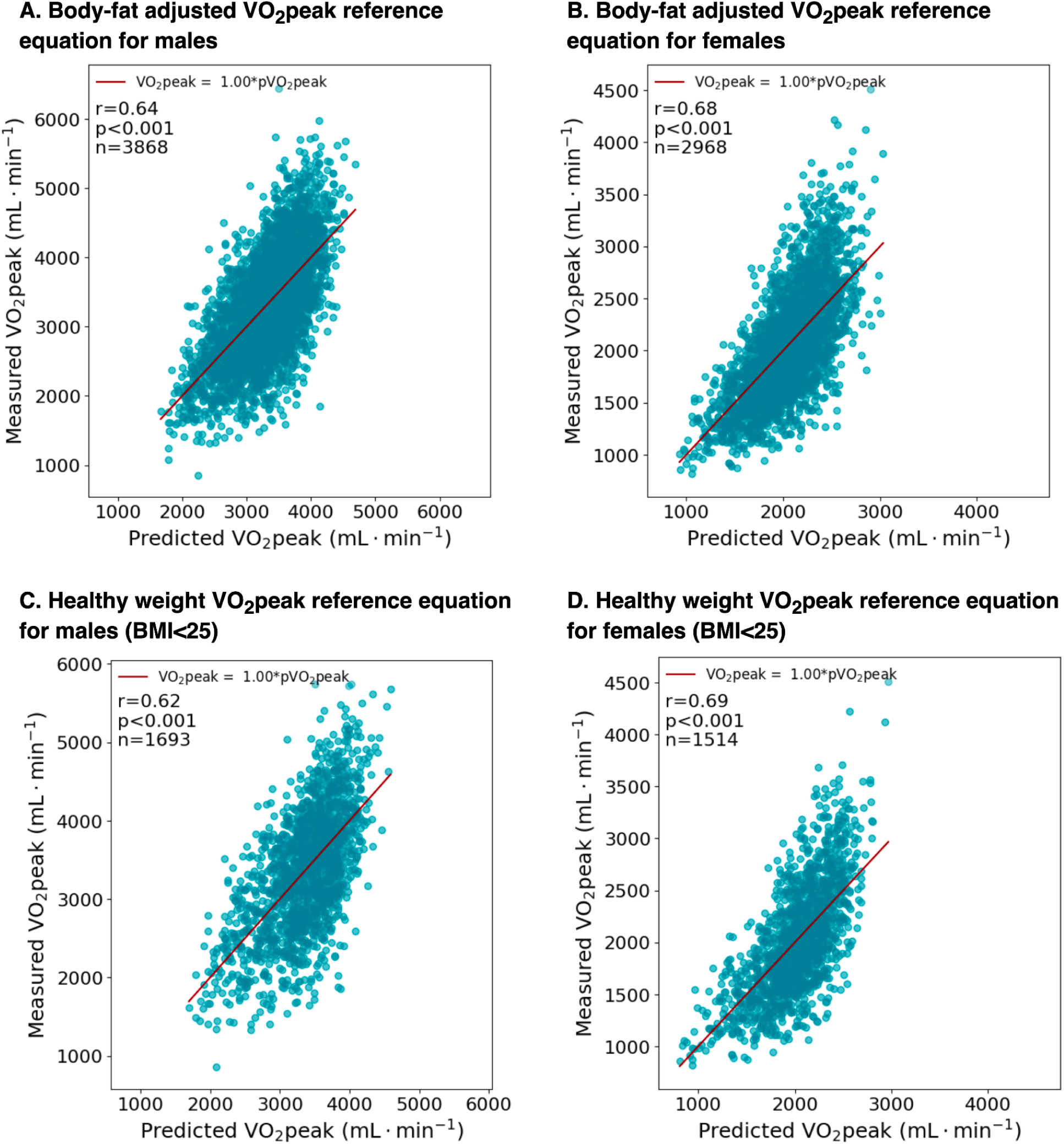
Measured versus predicted VO_2_peak using the simplified equations to mass and height terms.

**Supplemental Figure 2.**
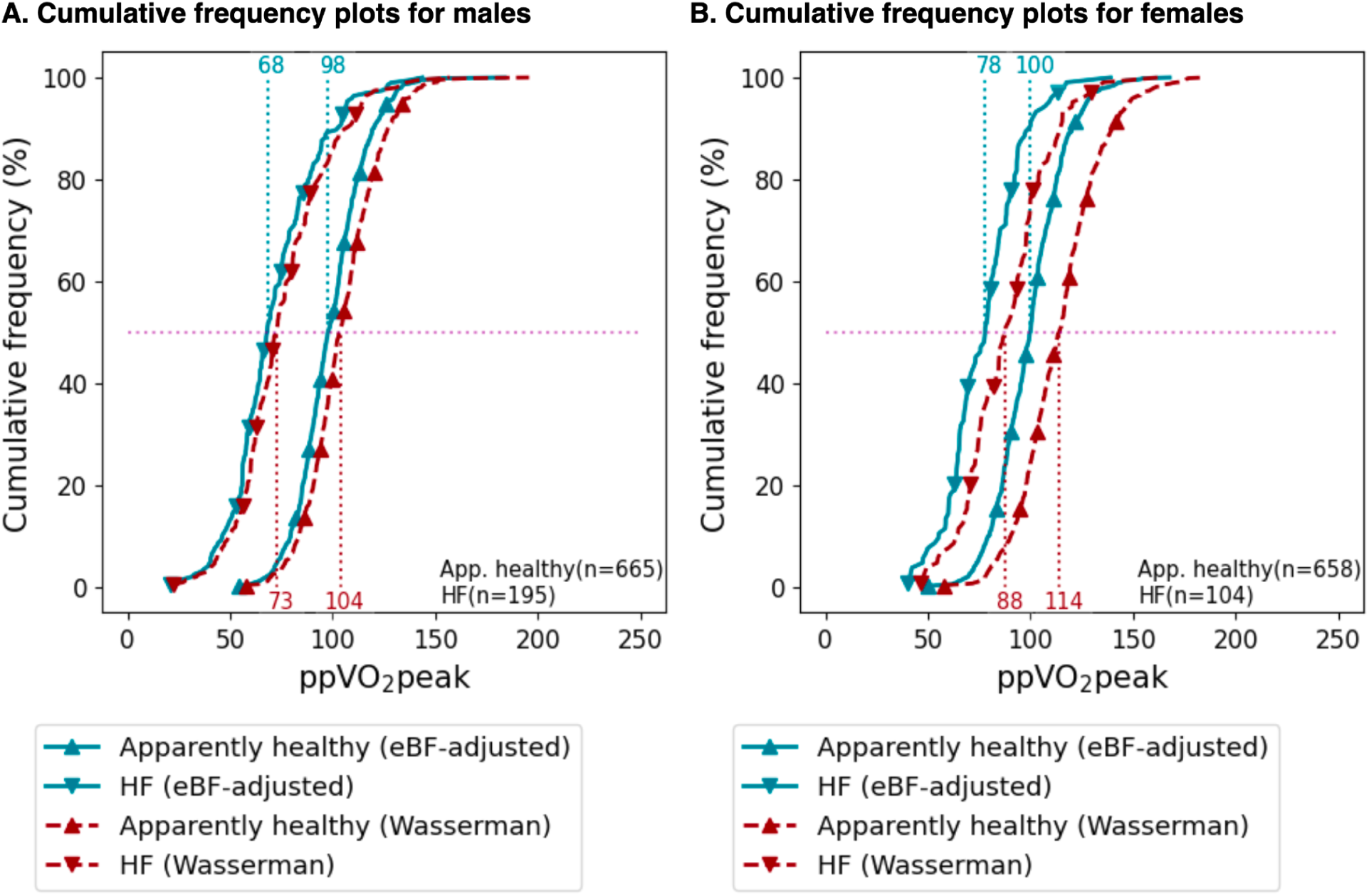
Application of body fat-adjusted equations in the obesity group.

---

**Supplemental Table 1.** Suggested percentage-predicted VO_2_peak thresholds for the novel equations and equivalence to the Wasserman equation.

|  | **FRIEND** | **Wasserman**<br>**(equivalent)** |
| --- | --- | --- |
| High | >135 | > 150 |
| Above average | 116-135 | 126-149 |
| Average | 85-115 | 92-125 |
| Below Average | 70-84 | 80-92 |
| Low | < 70 | < 80 |

